# The differential immune responses to COVID-19 in peripheral and lung revealed by single-cell RNA sequencing

**DOI:** 10.1101/2020.08.15.20175638

**Authors:** Gang Xu, Furong Qi, Hanjie Li, Qianting Yang, Haiyan Wang, Xin Wang, Xiaoju Liu, Juanjuan Zhao, Xuejiao Liao, Yang Liu, Ido Amit, Lei Liu, Shuye Zhang, Zheng Zhang

## Abstract

Understanding the mechanism that leads to immune dysfunction induced by SARS-CoV2 virus is crucial to develop treatment for severe COVID-19. Here, using single cell RNA-seq, we characterized the peripheral blood mononuclear cells (PBMC) from uninfected controls and COVID-19 patients, and cells in paired broncho-alveolar lavage fluid (BALF). We found a close association of decreased dendritic cells (DC) and increased monocytes resembling myeloid-derived suppressor cells (MDSC) which correlated with lymphopenia and inflammation in the blood of severe COVID-19 patients. Those MDSC-like monocytes were immune-paralyzed. In contrast, monocyte-macrophages in BALFs of COVID-19 patients produced massive amounts of cytokines and chemokines, but secreted little interferons. The frequencies of peripheral T cells and NK cells were significantly decreased in severe COVID-19 patients, especially for innate-like T and various CD8^+^ T cell subsets, compared to health controls. In contrast, the proportions of various activated CD4^+^ T cell subsets, including Th1, Th2 and Th17-like cells were increased and more clonally expanded in severe COVID-19 patients. Patients’ peripheral T cells showed no sign of exhaustion or augmented cell death, whereas T cells in BALFs produced higher levels of *IFNG, TNF, CCL4* and *CCL5* etc. Paired TCR tracking indicated abundant recruitment of peripheral T cells to the patients’ lung. Together, this study comprehensively depicts how the immune cell landscape is perturbed in severe COVID-19.

## Introduction

The coronavirus disease 2019 (COVID-19) caused by SARS-CoV-2 has been spreading rapidly worldwide, causing serious public health crisis. Although most SARS-CoV-2 infected cases have asymptomatic or mild-to-moderate diseases, around 10% of those infected may develop severe pneumonia and other associated organ malfunctions^1^. Old age, the male sex and underlying comorbidities are risk factors for causing severe COVID-19^2^, however, the pathogenesis mechanisms remains unclear. Immune perturbations were thought to play crucial roles in the COVID-19 pathogenesis. Indeed, many reports showed that lymphopenia and increased blood cytokine levels were closely associated with the development or recovery of severe COVID-19 and proposed to treat the patients by inhibiting cytokine storms^3–6^.

Recent studies have revealed more aspects of different immune players in COVID-19 infections. Although SARS-CoV-2 infection in host cells elicits robust secretion of chemokines and cytokines, it only weakly induces the IFNs^7^. In our earlier studies, we showed increased recruitment of highly inflammatory *FCN1*^+^ monocyte-derived macrophages in patients’ BALFs, suggesting their implication in the cytokine storm^8,9^. Intriguingly, T and B cell responses, and neutralizing antibodies recognizing SARS-CoV-2 were detected among most of the infected patients, with higher levels in those with old age and severe diseases^10–12^. The dysregulations of other immune cell compartments, such as monocytes, DCs, and innate-like T cells are also likely present in severe COVID-19^13–16^.

To unravel the barely known immune mechanisms underlying COVID-19 pathogenesis in an unbiased and comprehensive manner, we and others have applied the scRNA-seq to profile the immune cell heterogeneity and dynamics in BALFs, blood, and respiratory tract samples from the COVID-19 patients. Collectively, those studies revealed a stunted IFN response, depletion of NK and T lymphocytes, loss of MHC class II molecules and significantly elevated levels of chemokine production from monocytes in the patients^8,9,13,14,17–19^. However, a complete picture of the COVID-19 induced immune perturbation has not been generated. Here, we conducted scRNA-seq analysis of paired blood and BALF samples from the same COVID-19 patients. Our data revealed profound alterations of various immune compartments and depicted a dichotomy of peripheral immune paralysis and broncho-alveolar immune hyperactivation in COVID-19 patients.

## Results

### The overview of dysregulated peripheral immune landscape in severe COVID-19 patients

A high-quality single-cell RNAseq dataset composed of 200, 059 cells were generated that characterized peripheral immune cells from 3 healthy controls, 5 mild and 8 severe COVID-19 patients (Figure 1A). The metadata of these patients is listed in Table 1. Patients with mild diseases were all cured and discharged after 11–18 days hospitalization, 2 of the 8 patients with severe diseases succumbed, whereas other severe cases recovered after 12–58 days hospitalization. The clinical course of this patient cohort is similar to earlier reports, where elderly patients with underlying diseases were prone to develop severe symptoms and showed higher mortality^1,2,20^. In addition, the plasma levels of IL-6 and C-reactive proteins (CRP) in severe patients were higher, while lymphocyte counts were reduced, indications of both cytokine storm and lymphopenia.

**Fig 1.**
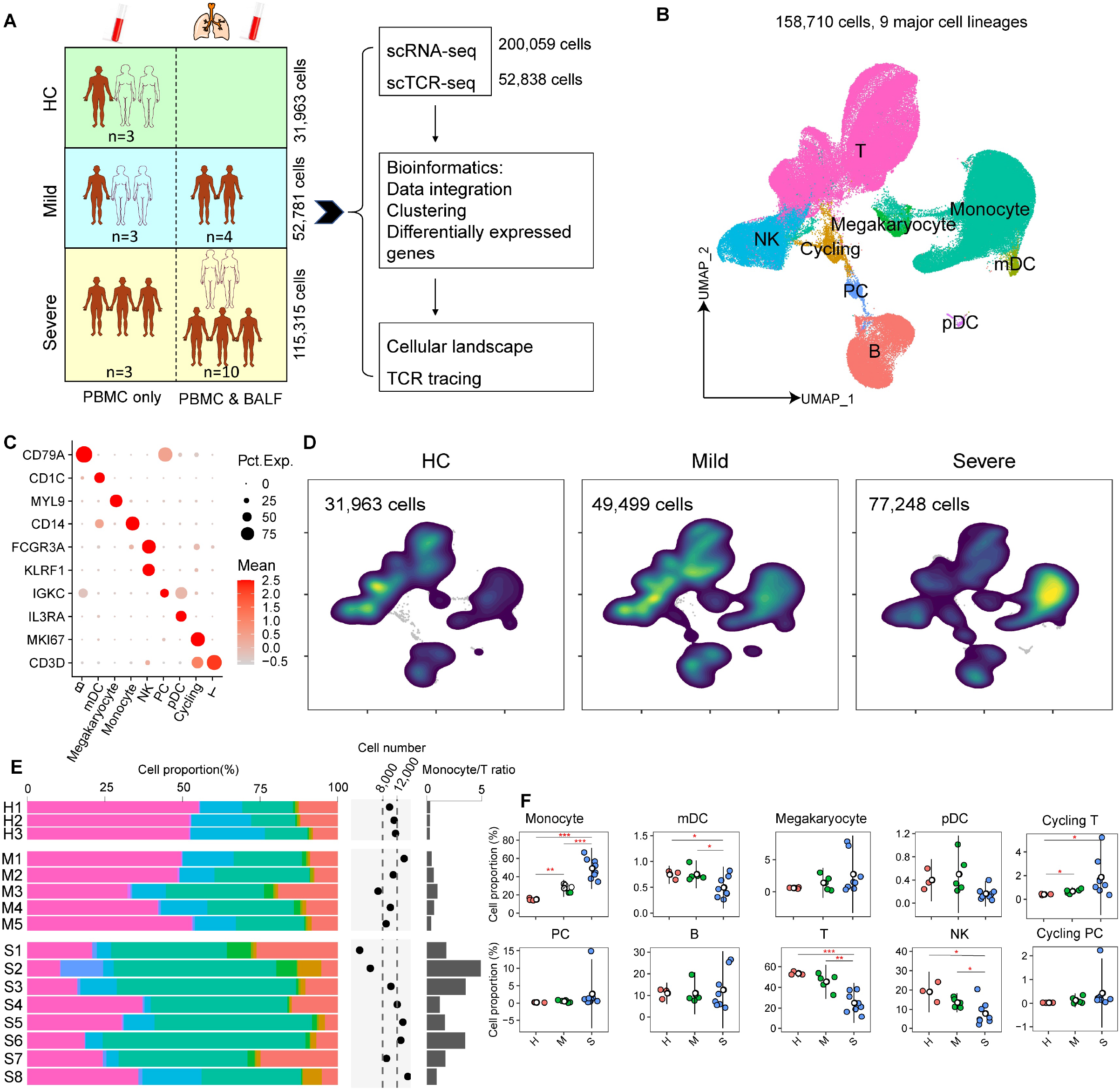
<u>Single-cell analysis of peripheral blood mononuclear cells from patients with COVID-19</u>. (A) The carton outlines the study design. PBMC and BALF cells from COVID-19 patients and healthy controls were collected for scRNA-seq characterization using the 10x Genomics platform. Number of samples, analyzed cells, and samples with PBMC and BALF cells simultaneously collected from the same patient are indicated. (B) The UMAP projection of the combined PBMCs scRNA-seq dataset, identifies 9 major cell types. PC stands for plasma cells. (C) The specific markers for identifying each immune cell types in (B) are indicated. (D) Density plots shows the UMAP projection of PBMCs from COVID-19 patients and controls. (E) The bar plot shows the proportions of each cell types in PBMCs from individual subjects. The cell numbers and ratios of monocyte / T-cells are listed to the right-side. (F) Comparisons of percentages of each cell types in PBMCs (Cycling cells were re-clustered into T and PC subsets) between the two COVID-19 groups and controls (two-sided Student’s *t*-test, *p<0.05, **p<0.01, ***p<0.001.

**Table 1.** Clinical data of the enrolled COVID-19 patients and healthy controls examined by scRNA-seq

| Sample NO. | S1 | S2 | S3 | S4 | S5 | S6 | S7 | S8 | M1 | M2 | M3 | M4 | M5 | H1 | H2 | H3 |
| --- | --- | --- | --- | --- | --- | --- | --- | --- | --- | --- | --- | --- | --- | --- | --- | --- |
| Severity | severe | severe | severe | severe | severe | severe | severe | severe | mild | mild | mild | mild | mild | HC | HC | HC |
| Age | 73 | 46 | 67 | 65 | 62 | 57 | 63 | 66 | 35 | 36 | 42 | 49 | 32 | 66 | 36 | 31 |
| Gender | Male | Male | Male | Female | Male | Female | Male | Male | Male | Male | Male | Female | Female | Female | Female | Male |
| Symptom onset day | 2020/1/20 | 2020/1/21 | 2020/1/17 | 2020/1/4 | 2020/1/11 | 2020/1/21 | 2020/1/8 | 2020/1/3 | 2020/1/9 | 2020/1/9 | 2020/1/18 | 2020/1/22 | 2020/1/21 |  |  |  |
| First symptom | Fever | Fever/Cough | Fever/diarrhea | Fever/Cough | Fever/Cough | Cough | Fever/Cough | Fever | Cough | Cough | Fever/Cough | Fever/Cough | Fever |  |  |  |
| Hospitalization date | 2020/1/22 | 2020/1/22 | 2020/1/22 | 2020/1/20 | 2020/1/15 | 2020/1/23 | 2020/1/15 | 2020/1/11 | 2020/1/15 | 2020/1/16 | 2020/1/21 | 2020/1/23 | 2020/1/23 |  |  |  |
| Chronic basic disease | none | none | Bronchitis | diabetes | none | none | COPD | hyperpression | none | none | None | None |  |  |  |  |
| CoV-RNA | + | + | + | + | + | + | + | + | + | + | - | - | - | + | - | + |
| Influenza A virus | - | - | - | - | - | - | - | - | - | - | - | - | - | - | - | - |
| Influenza B virus | - | - | - | - | - | - | - | - | - | - | - | - | - | - | - | - |
| Respiratory syncytial virus | - | - | - | - | - | - | - | - | - | - | - | - | - | - | - | - |
| Adenovirus | - | - | - | - | - | - | - | - | - | - | - | - | - | - | - | - |
| CT finding | Bilateral pneumonia | Unilateral pneumonia | Bilateral pneumonia | Unilateral pneumonia | Bilateral pneumonia | Bilateral pneumonia | Bilateral pneumonia | Bilaeral pneumonia | Bilateral pneumonia | Bilateral pneumonia | Bilaeral pneumonia | Bilaeral pneumonia | Unilateral pneumonia |  |  |  |
| PBMC sampling date | 2020/2/2 | 2020/2/2 | 2020/1/28 | 2020/1/28 | 2020/1/30 | 2020/1/29 | 2020/1/29 | 2020/1/31 | 2020/1/29 | 2020/1/22 | 2020/1/23 | 2020/1/28 | 2020/1/28 |  |  |  |
| Outcome-date | Cure/2020/2/20 | Cure/2020/3/8 | Cure/2020/3/20 | Cure/2020/3/9 | Cure/2020/2/27 | Cure/2020/3/7 | Dead/2020/2/16 | Dead/2020/2/16 | Cure/2020/2/3 | Cure/2020/1/27 | Cure/2020/2/3 | Cure/2020/2/8 | Cure/2020/2/9 |  |  |  |
| IL6 (pg/ml) | 35.54 | 17.99 | 36.14 | 14.61 | 69.18 | 22.87 | 225.3 | 353.2 | 4.43 | 1.92 | 18.33 | 35.2 | 4.59 |  |  |  |
| CRP (pg/ml) | 6.94 | 50.63 | 112.78 | 2.4 | 88.88 | 7.86 | 33.11 | 189.23 | 6.75 | 1.31 | 12.75 | 60.01 | 8.63 |  |  |  |
| WBC(10<sup>9</sup>/L) | 5.75 | 12.42 | 11.47 | 9.59 | 7.13 | 32.04 | 11.47 | 7.24 | 5.38 |  | 4.13 | 5.67 | 4.55 |  |  |  |
| NEUT(10<sup>9</sup>/L) | 5.49 | 11.5 | 10.52 | 8.62 | 5.45 | 29.19 | 10.11 | 5.56 | 2.64 |  | 2.58 | 3.77 | 3.02 |  |  |  |
| LYMPH(10<sup>9</sup>/L) | 0.16 | 0.33 | 0.46 | 0.59 | 0.82 | 1.45 | 0.88 | 0.78 | 2.14 |  | 1.09 | 1.19 | 1.09 |  |  |  |
| MONO(10<sup>9</sup>/L) | 0.1 | 0.58 | 0.47 | 0.37 | 0.79 | 1.39 | 0.45 | 0.18 | 0.44 |  | 0.46 | 0.59 | 0.31 |  |  |  |
| NEUT/LYMPH | 34.3 | 34.8 | 22.9 | 14.6 | 6.6 | 20.1 | 11.5 | 7.1 | 1.2 |  | 2.4 | 3.2 | 2.8 |  |  |  |
| CD3 (cells/ul) | 66 | 92 | 141 | 201 | 427 | 744 | 268 | 629 | 1782 |  | 527 | 1258 | 1261 |  |  |  |
| CD4 (cells/ul) | 59 | 58 | 93 | 149 | 354 | 563 | 181 | 495 | 1141 |  | 301 | 684 | 671 |  |  |  |
| CD8 (cells/ul) | 5 | 30 | 37 | 45 | 66 | 166 | 80 | 128 | 566 |  | 204 | 420 | 565 |  |  |  |

The clustering analysis showed 29 clusters and 10 major cell types annotated by marker genes, including T cell (*CD3D*), natural killer (NK) cell (*KLRF1*), B cell (*CD79A*), monocyte (*CD14, FEGR3A*), myeloid dendritic cells (mDC) (*CD1C*), plasmacytoid dendritic cells (pDC) (*IL3RA*), and plasma cells (PC) (*IGKC*), megakaryocyte (*MYL9*), cycling cells (*MKI67*) and erythrocyte (*HBB)* (Figure 1B-C, Figure S1A-S1B). By visual inspection, the data integration was efficient and showed no significant batch-effect (Figure S1C). Erythrocytes were not included in subsequent analysis and cycling cells were reclustered into cycling T, cycling PC and cycling NK cells based on specific markers (Figure S1D-S1E). This dataset indicated significantly dysregulated peripheral immune landscapes in COVID-19 patients compared to health controls, especially among those severe cases (Figure 1D). The most prominent changes included an expansion of monocyte and cycling T cells, and a reduction of NK, T and mDC populations, thus resulting in largely increased monocyte/T-cell ratios in COVID-19 patients (Figure 1E-1F). The frequency of pDC was also decreased in severe COVID-19, although the difference was not statistically significant. Together, these data show that SARS-CoV-2 infection greatly perturbs the blood immune cell compartments, particularly in those with severer diseases.

### Remodeling of circulating myeloid cell populations in patients with COVID-19

We observed that the proportions of monocytes were increased in COVID-19 patients, especially in those with severe diseases. To further understand the remodeling of myeloid cell compartment, we reclustered myeloid cells and identified five distinct cell types including CD14^+^ classic monocyte, CD14^+^CD16^+^ intermediate monocytes, CD16^+^ non-classic monocytes, DC1 and DC2 (Figure 2A and Figure S2A). The composition of myeloid cells in severe COVID-19 patients differed significantly from that of mild cases and controls. Proportion of CD14^+^ monocyte increased significantly in severe COVID-19 compared to that in mild COVID-19 and control group, whereas those of CD16^+^ non-classical monocyte (vs. controls), CD14^+^CD16^+^ monocytes (vs. mild COVID-19) and DC2 significantly (vs. mild COVID-19 & controls) decreased in severe COVID-19 (Figure 2B-2C).

**Fig 2.**
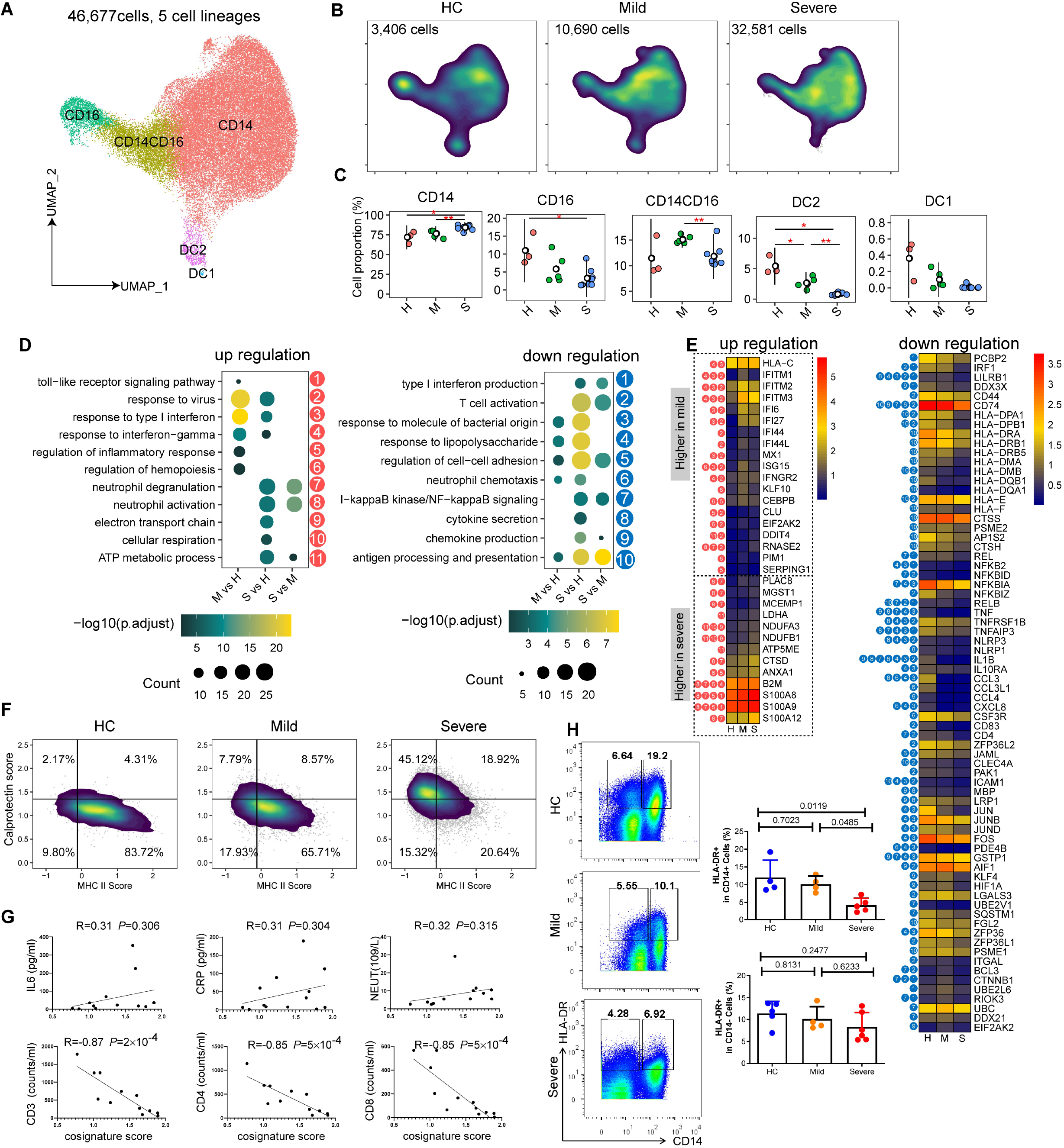
<u>Single-cell analysis of peripheral myeloid cell compartments in patients with COVID-19</u>. (A) UMAP plot of the five types of myeloid cells in PBMCs. (B) Density plots show the UMAP projection of peripheral myeloid cells from COVID-19 patients and controls. (C) Comparisons of percentages of each peripheral myeloid cell types between the two COVID-19 groups and controls (two-sided Student’s *t*-test, *p<0.05, **p<0.01, ***p<0.001). (D) Enrichment of GO biological process (BP) terms for up-regulated genes (left) and down-regulated genes (right) in blood CD14^+^ monocyte comparisons between mild and HC (M vs H), severe and HC (S vs H), and severe and mild groups (S vs M). (representative terms are shown, adjusted p < 0.01 as indicated by the colored bar). (E) The heatmaps show the selected differentially expressed genes and their associated GO terms as indicated in (D). (Natural logarithm fold change > 0.41 or < –0.41, adjusted p < 0.01). (F) Density plots show the composite MHC class II signature scores and calprotectin signature scores of peripheral CD14^+^ monocytes in 2D maps. The horizontal and vertical lines separating the four quadrants represent the median scores of all CD14^+^ monocytes. The percentages of cells in each quadrant are indicated. (G) The Pearson correlation of “MDSC-like signature score” and plasma CRP, IL-6 levels, blood neutrophil, CD3^+^, CD4^+^ and CD8^+^ T cell counts. (H) Left panel shows the representative flow cytometric data of HLA-DR expression on CD14^+^ and CD14^−^ PBMCs. Right plot shows the summarized data from more subjects (two-sided Student’s *t*-test).

CD14^+^ monocytes represent the major peripheral myeloid cell type and differential UMAP projection patterns of CD14^+^ monocytes between COVID-19 and controls (Figure 2B) indicated perturbed transcriptome features. Among the differentially expressed genes (DEGs) in CD14^+^ monocytes, we found 116 and 134 upregulated genes in mild COVID-19 and severe COVID-19 cases compared to controls, versus only 74 upregulated genes in comparison between two COVID-19 groups. In contrast, we found 217 and 160 downregulated genes in severe COVID-19 compared to controls or mild cases respectively, versus only 104 downregulated genes in comparison between mild COVID-19 and controls (Figure S2B and Table S1. GO terms of upregulated DEGs included response to virus, type I IFN and IFN-γ in both COVID-19 groups, and neutrophil activation and energy metabolism pathways in severe COVID-19 (Figure 2D). Unexpectedly, GO analysis of downregulated DEGs indicated deficient monocyte functions mainly in severe COVID-19 cases, such as decreased type I IFN production, cytokine secretion, chemokine production and antigen processing and presentation (Figure 2D). We further examined the DEGs associated with those GO terms. Indeed, many canonical interferon-stimulating genes (ISGs), including *ISG15, IFITM1, IFITM3, MX1, IRF7, IFI27 etc*., were expressed at higher levels in COVID-19 patients than controls, while the genes associated with neutrophil activation, including *S100A8, S100A9, S100A12, CLU and RNASE2 etc*., were expressed at higher levels in severe COVID-19 than mild COVID-19 and controls (Figure 2E). Despite of upregulated ISGs, we failed to detect higher production of type I or type III IFNs in COVID-19 than controls. Regarding the downregulated geneset, MHC class II molecules, including *HLA-DQA1, HLD-DRA, HLA-DRB1, HLA-DMB, HLA-DMA, etc*., and those cytokine/chemokine genes, including *IL1B*, *TNF, CCL3, CCL4 and CXCL8* were expressed at lower levels in COVID-19 patients, especially those with severe diseases (Figure 2E). Thus, those upregulated DEGs in CD14^+^ monocytes from COVID-19 patients reflect the immune response to SARS-CoV-2 infection, while the downregulated DEGs in CD14^+^ monocytes from patients with severe COVID-19 suggest an immune paralyzed status of those cells.

MDSCs are a population of heterogeneous myeloid cells expanded during inflammatory conditions and could suppress T-cell responses^21,22^. In peripheral blood, monocytic MDSCs have the phenotype CD14^+^ HLA-DR^−/lo^, whereas monocytes are HLA-DR^+^^23,24^. Downregulation of MHC class II molecules, increased calprotectin proteins like S100A8 and S100A9, and immune suppressive functions are reported features of MDSCs. Indeed, by their unique composite scores of MHC class II molecules (lower levels) and S100A8 family molecules (higher levels) versus those scores in mild COVID-19 and controls, we identified that the monocytes in severe COVID-19 highly resembled MDSCs (Figure 2F). Decreased levels of HLA-DR in CD14^+^ monocyte from severe patients were further validated by flow cytometry (Figure 2G) and also reported by other studies^14,18^. Intriguingly, MDSCs-like scores in our study positively correlate with serum CRP, IL-6 levels and neutrophil-to-lymphocyte ratio, and negatively correlate with decreased blood CD3^+^, CD4^+^ and CD8^+^ T cell counts (Figure 2H).

Together, our scRNA-seq characterization revealed a multifaceted remodeling of peripheral myeloid compartment in COVID-19 patients. While the circulating monocytes in COVID-19 patients are featured by heightened ISG responses, they produce little interferons, cytokines and chemokines. Furthermore, the combined loss of dendritic cells and emergence of monocytic MDSCs suggest their involvements in the immune paralysis of severe COVID-19 patients.

### Abnormally activated lung monocyte-macrophages in severe COVID-19

Recently, we discovered the aberrant activation of BALF monocyte-macrophages in severe COVID-19^8,9^. Here, to further understand the connection between the lung monocyte-macrophages and their blood counterparts and assess their differential roles in COVID-19, we studied paired BALF and blood samples from 2 mild and 5 severe patients. The integration analysis of BALF and circulating myeloid cells showed clusters of neutrophil (*FCGR3B*), mDC (*CD1C*), monocyte-macrophages (*CD14, FCGR3A and CD68*) (Figure 3A and Figure S3A). Macrophage subset classification markers, including *FCN1, SPP1* and *FABP4*, were differentially expressed by circulating and BALF monocyte-macrophages from patients with mild or severe COVID-19(Figure 3B). Analysis of differentiation trajectory of circulating and BALF monocyte-macrophages from the same patient revealed a consensus blood-toward-BALF course (Figure 3C and Figure S3B), consistent with the recruitment to peripheral monocytes into inflammatory tissues as expected.

**Fig 3.**
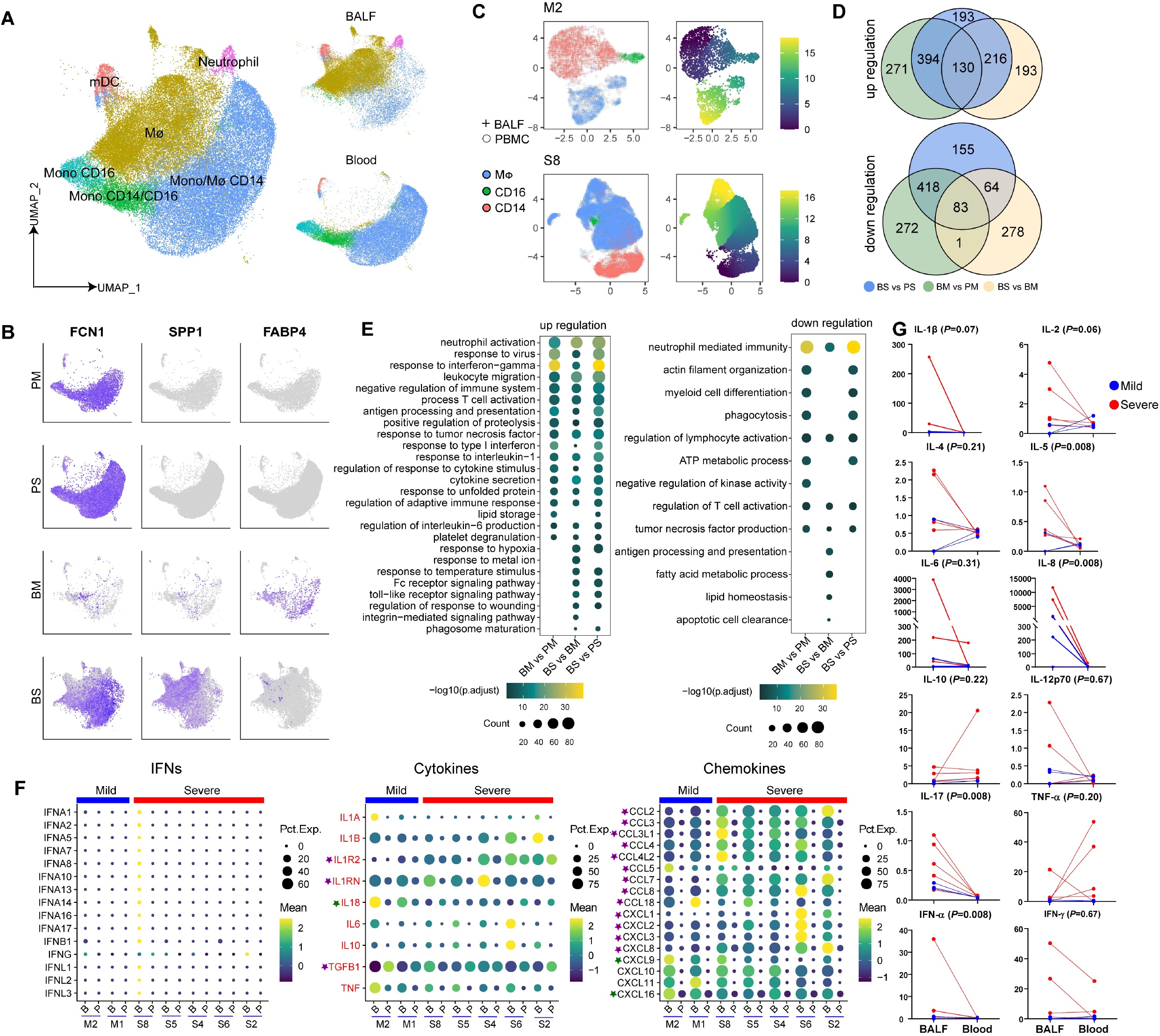
<u>Abnormally activated lung monocyte-macrophages in severe COVID-19</u>. (A) The myeloid cell data from 2 mild and 5 severe COVID-19 patients who had paired PBMC and BALF samples, were integrated and presented on the UMAP. (B) The expression of monocyte-macrophage markers *FCN1, SPP1* and *FABP4* were projected to UMAP from (A). (C) Differentiation trajectory of the blood monocytes and BALF monocyte-macrophages from two representative COVID-19 patients, analyzed independently. (D) Venn diagram shows the number of up-regulated and down-regulated DEGs in monocyte-macrophages comparisons as indicated. BM: BALF of mild cases; BS: BALF of severe cases; PM: Peripheral cells of mild cases; PS: Peripheral cells of severe cases. (Natural logarithm fold change > 0.41 or < –0.41, adjust p < 0.01) (E) Enrichment of GO biological process (BP) terms for up-regulated genes (left) and down-regulated genes (right) in monocyte-macrophage comparisons as indicated. (Selected terms are shown, adjusted p value is indicated by the colored bar). (F) Heatmaps show the expression of selected interferon, cytokine and chemokine genes in paired blood and BALF monocyte-macrophages derived from the same patient. Stars indicate that the genes are differentially expressed in BALF monocyte-macrophages between mild and severe COVID-19. Purple and green stars show the gene expression are significantly upregulated in severe COVID-19 and mild COVID-19 group respectively (MAST; p < 0.01). (G) The levels of selected cytokines and chemokines in paired BALF and plasma samples were measured by CBA (two-sided Wilcoxon test between BALF and PBMC of severe patients).

Next, we performed transcriptome analysis of circulating and BALF monocyte-macrophages to understand their functional status. Among the DEGs, there were 524 shared upregulated genes and 501 downregulated genes in BALF monocyte-macrophages versus those in blood, identified from both mild and severe COVID-19 patients (Figure 3D and Table S2). Such a large number of DEGs suggested significant difference existed between the peripheral and lung monocyte-macrophages. Indeed, the GO analysis revealed broad activation of multiple immune pathways in BALF monocyte-macrophages, including response to IFNs and cytokines, neutrophil activation and leukocyte migration, while the pathway involved in myeloid cell differentiation, ATP metabolism *etc*. were enriched in blood monocytes (Figure 3E). In addition, these comparisons revealed perturbed pathways in BALF monocyte-macrophages relevant to severe COVID-19. e.g. responses to hypoxia, high temperature, metal ion, wounding and Fc receptor signaling pathways were specially upregulated (Figure 3E), while pathways related to alveoli macrophage functions were downregulated, including lipid metabolism, apoptotic cell clearance and antigen presentation (Figure 3E). The representative DEGs involved in those pathways were shown in Figure S3C.

Monocyte-macrophages were thought to play key roles in driving the cytokine storm underlying the development of severe COVID-19^25^. Therefore, we examined the cytokine and chemokine levels in monocyte-macrophages in paired blood and BALF samples from the same patient. We found that all types of IFNs (*IFNAs, IFNB, IFNG* and *IFNLs*) were minimally expressed by monocyte-macrophages, whereas cytokines (*IL1A, IL1B, IL1R2, IL1RN, IL18, IL6, TNF, IL10* and *TGFB1*) and multiple chemokines were highly expressed in monocyte-macrophages from BALFs but not in those from paired blood samples (Figure 3F). Intriguingly, we observed significantly higher levels of anti-inflammatory cytokines (*IL1R2, IL1RN* and *TGFB1*) and lower levels of *IL18* in BALF cells from severe COVID-19 than mild cases, whereas classical pro-inflammatory cytokines (*IL1A, IL1B, IL6* and *TNF*) were comparable between the two groups (Figure 3F). In contrast, as shown in our earlier studies, monocytes- and neutrophils-recruiting chemokines (*CCL2, CCL3, CCL4, CCL7, CCL8, CXCL1, CXCL2, CXCL3* and *CXCL8*) recruiting monocytes and neutrophils were highly expressed, whereas T cell recruiting chemokines (*CXCL9* and *CXCL16*) recruiting T cells were less expressed by monocyte-macrophages in BALFs of severe COVID-19 monocyte-macrophages than those in mild cases (Figure 3F). The higher levels of cytokines (IL-1β and IL-6 *etc*.) and IL-8 in BALFs than paired plasma was further validated at the protein levels, particularly exemplified by the extremely high levels of IL-8 in BALFs (Figure 3G). Thus, these paired analyses revealed a restricted involvement of tissue monocyte-macrophages in cytokine storms during severe COVID-19, through producing chemokines and recruiting more monocytes and neutrophils, but unlikely attribute to increased production of pro-inflammatory cytokines.

### The dysregulated peripheral T cell compartments in COVID-19 patients

NK and T lymphocytes are important anti-viral immune cells, which are depleted in severe COVID-19^13,26^. To further understand the dysregulated NK and T cell compartments, we re-clustered those cells and identified 18 subsets (Figure 4A and Figure S4A). NK cells highly expressed *KLRF1, KLRC1* and *KLRD1*. Cycling T cells expressed *MKI67*. Innate-like T cells included MAIT (*SLC4A10*), γδ T (*TRGV9*) and NKT cells (*CD3E, KLRF1*). CD4^+^ T cells included CD4-Naive (*CCR7, SELL*), CD4-*LTB*, CD4-*GZMK*, CD4-*GATA3*, CD4-*CCR6*, CD4-*ICOS*, CD4-*GZMB*, Treg-*SELL* and Treg-*CTLA4* subsets, whereas CD8^+^ T cells included CD8-Naive (*CCR7, SELL*), CD8-*LTB*, CD8-*GZMK* and CD8-*GZMB* subsets. Pseudo-time trajectory analysis was performed to infer lineage relationship among those CD4^+^ and CD8^+^ T cells subsets. Paired TCR clonotype analysis revealed increased clonal expansion along the inferred trajectories (Figure 4B).

**Fig 4.**
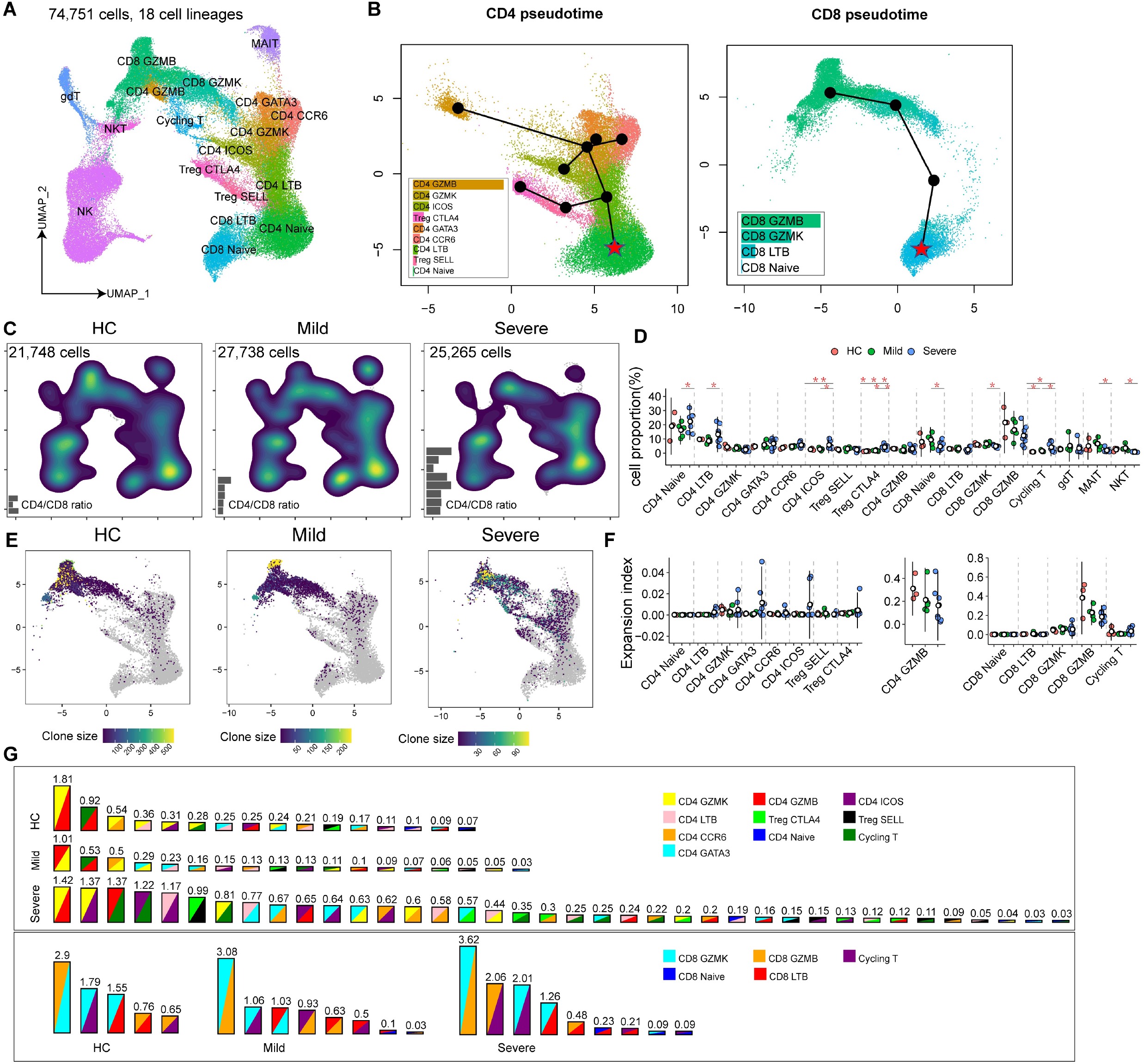
<u>Single-cell analysis of peripheral NK and T cell compartments in patients with COVID-19</u>. (A) UMAP plot of the 18 subsets of NK and T cells in PBMCs. (B) Pseudo-time differentiation trajectory of the peripheral CD4^+^ and CD8^+^ T cells subsets performed by slingshot. The bar plots in the corner shows the percentages of clonally expanded cells in each T cell subsets. (C) Density plots show the UMAP projection of peripheral NK and T cells from COVID-19 patients and controls. (D) Comparisons of percentages of each peripheral NK and T cell types between the two COVID-19 groups and controls (two-sided Student’s *t*-test, *p<0.05, **p<0.01, ***p<0.001). (E) Clonally expanded T cells from COVID-19 patients and controls are projected into UMAP from (A). (F) Clonal expansion indexes of T cell subsets from COVID-19 patients and controls, are separately displayed. (G) T cell state transition status among any two clusters is inferred by their shared TCR clonotypes. Each T cell cluster is represented by a unique color. The numbers above the bar indicate the percentages of cells sharing TCRs in those two clusters.

Cell density UMAP projections revealed an obviously perturbed T cell landscape in severe COVID-19 compared to mild COVID and control groups (Figure 4C). Percentages of innate-like T cells, including MAIT and NKT cells were significantly lower in severe COVID-19 than those in mild COVID-19. Percentages of CD8-Naive, CD8-GZMK and CD8-GZMB subsets were also lower in severe COVID-19 patients than those in mild cases, although the difference in CD8-GZMB comparison was not statistically significant (Figure 4D). In contrast, the percentages of several CD4^+^ T cell subsets, including CD4-Naive, CD4-LTB, CD4-ICOS, Treg-CTLA4, as well as cycling T cells, were significantly increased in severe COVID-19 than those in mild COVID-19. The percentages of CD4-GATA3 and CD4-CCR6 also showed an increasing trend in severe COVID-19 (Figure 4D). In addition, sc-TCR analysis revealed increased clonal expansion levels in several CD4^+^ but not CD8^+^ T cell subsets in severe COVID-19 than those in mild cases (Figure 4E-4F). Consistently, TCR sharing analysis among different T cell subsets revealed much more active interchange between different CD4^+^ T cell subsets and cycling T cells, but not among CD8^+^ T cell subsets during severe COVID-19 (Figure 4G). Together, our data revealed the preferential activation of CD4^+^ T cell responses but significant depletion of multiple innate-like T cell and CD8^+^ T cell subsets in peripheral blood as the featured T cell perturbations in severe COVID-19. To further explore the clues of CD8^+^ T cell lymphopenia, we conducted transcriptome comparisons of MAIT, CD8-GZMK and CD8-GZMB subsets between the patients and control groups (Figure S4C and Table S3). There was no evidence of T cell exhaustion, activation of cell death pathways and cytokine productions in those cells from COVID-19 patients, although the pathways related to virus infection and IFNs responses were identified (Figure S4D). Similar findings were also noticed by other groups^16^; thus, exhaustion and cell death are unlikely the major causes for T cell loss during COVID-19.

### Tracking T cell function and migration across peripheral blood and BALFs

We sought to study T cell movement from blood to BALFs in paired samples from COVID-19 patients. First, we integrated data of NK & T cells from PBMC and BALF. Cells were re-clustered into 9 major types, including NK cells, MAIT, CD4 Naive, CD4 Tm, Treg, CD8 Naive, CD8 Tm, CD8 IL7R and Cycling T cells (Figure 5A and Figure S5A). PBMC included plenty of naive CD4^+^ and CD8^+^ T cells, in contrast there were fewer naive T cells in BALFs, which were mainly composed of NK cells, CD4 Tm, CD8 Tm and cycling T cells (Figure 5B). We performed gene expression analysis to determine the functional divergence of NK and T cells across the peripheral blood and BALFs (Table S4. Compared with peripheral counterparts, we observed that response to virus, type I IFN and IFN-γ were commonly activated in NK, CD4 Tm and CD8 Tm cells from BALFs (Figure S5B). We also noticed higher levels of cytokines including *IFNG, TNF, CSF1, TNFSF10* and *TNFSF13B*; chemokines including *CCL3, CCL4* and *CCL5*; IL-15 signaling modules including *IL15RA, IL2RB, IL2RG, JAK1* and *STAT3* in BALF cells (Figure 5C). However, the levels of other T cell relevant cytokines, including *IL4, IL5, IL10, IL13, IL17A, IL17F, IL21, IL25* and *IL33* were not detected in either blood or BALFs (Figure S5C).

**Fig 5.**
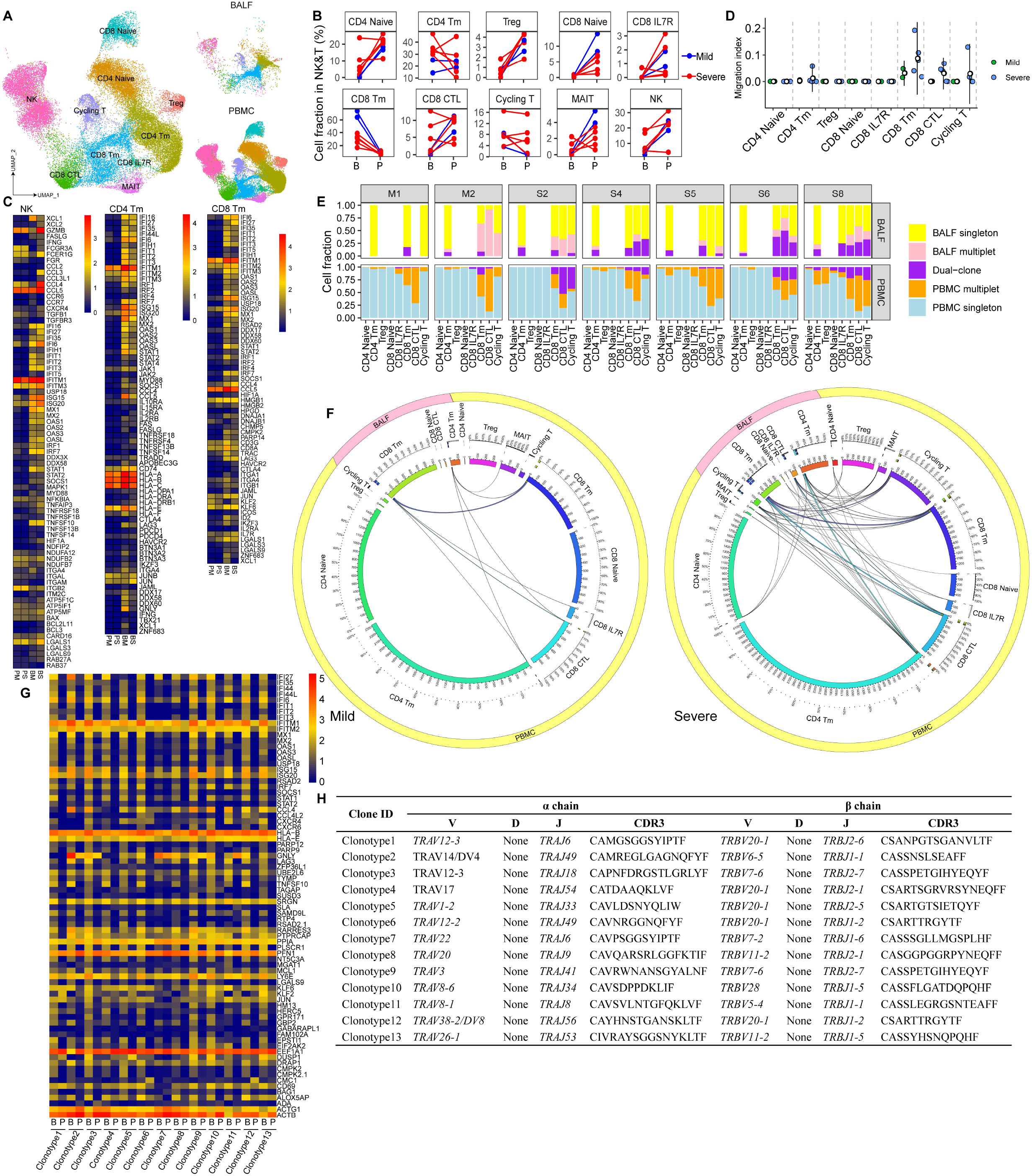
<u>Tracking T cells across peripheral blood and BALFs in patients with COVID-19</u>. (A) The T cell data from 2 mild and 5 severe COVID-19 patients who had paired PBMC and BALF samples, were integrated and presented on the UMAP. (B) The percentages of each T cell subset in paired BALF (B) and PBMC (P) of the same patient are compared. (C) Heatmaps display the selected DEGs in NK, CD4 Tm and CD8 Tm cells in BALF (B) or PBMC (P) from the mild (M) or severe (S) COVID-19 patients. (Natural logarithm fold change > 0.41,, adjust p < 0.01). (D) The migration index in each T cell subset across paired PBMC and BALF from seven patients were shown (STARTRAC-migr indices). (E) TCR clonotypes were classified into five different types as indicated by different color bars (singleton indicates non-expanded TCR clonotype, multiplet indicates expanded TCR clonotype, dual-clone indicates those clonotype shared in paired PBMC and BALF samples). The bar plots show the percentages of different types of TCR clonotypes in different T cell subsets from paired PBMC and BALF samples. (F) The circus plot shows the degree of TCR clonotype sharing across different T cell subsets in PBMC and BALF from mild and severe COVID-19 groups. (G) Heatmap shows the selected DEGs in each T cell clones derived from the top 13 TCR clonotypes shared across PBMC versus BALF compartments. (Natural logarithm fold change > 0.41, adjust p < 0.01). (H) The V, J genes of the TCR *α* and *β* chains of the top 13 dual clonotypes are listed, the amino acid sequences of their CDR3 are shown.

Next, we utilized the TCR clonotype information to track the migrating T cells in blood and BALF compartment. The T cell clonal expansion status and clonotype sharing were assessed in patients with paired blood and BALF samples (Figure 5D-5E). Interestingly, considerable proportion of BALF T cells belonging to CD8 Tm, CD8 CTL and cycling T cells subsets, could trace their clonotypes back to the paired blood counterparts, especially in those patients with severe COVID-19 (Figure 5F). Here, only two mild cases (M1 and M2) had paired blood and BALF samples for the analysis, and it showed a lower degree of clonotype sharing across the two compartments (Figure 5F). The higher levels of chemokines from the lung of severe COVID-19 is likely to lead to increased T cell infiltration. To assess the functional adaptation of migrating T cells, we performed transcriptional analysis between blood vs. BALF T cells derived from the same T-cell clonotypes (Figure 5H). We found increased expression of ISGs, *CCL4, CXCR4, CXCR6, CD69* and *GNLY etc*. in BALF cells than those in blood, consistent with an activated and tissue resident phenotype. (Figure 5G and Table S5) Together, these data revealed the active recruitment and peculiar activation of peripheral T cells in human lungs infected by SARS-CoV-2.

## Discussion

The scRNA-seq has been recently been applied to study host immune response in COVID-19 by us and others^8,9,13,17–19^. Although these studies helped to reveal several aspects of the COVID-19 pathogenesis, a complete picture has yet been generated. Here, we integrated scRNA-seq analysis of paired blood and BALFs, depicted unprecedented details about the altered immune cell landscape in COVID-19 patients, increasing our mechanistic and systemic understanding of COVID-19 immunopathogenesis, such as cytokine storm and lymphopenia.

IFN production is stunted in SARS-CoV-2 infected cells and COVID-19 patients^7,27^. Consistent with these earlier reports, here we found that although ISGs in COVID-19 patients were induced in monocyte-macrophages, neither type I or type III IFNs were produced. However, the higher levels of ISGs in monocyte-macrophages and T cells from mild cases than those in severe COVID-19, still supported a heightened IFN responses linked to resolving the diseases. It is unclear what causes the imbalance of inflammation and IFN production in COVID-19, we assume that the loss of pDCs in severe cases may partly contribute to the diminished IFN production. Notably, in contrast to their blood counterparts, monocyte-macrophages in paired BALFs produced extremely higher levels of cytokines and chemokines, especially from those severe COVID-19 patients. Thus, massive tissue-resident production of cytokines / chemokines and lack of IFN induction suggest a crucial role played by local but not circulating monocyte-macrophages in fueling the cytokine storm during severe COVID-19.

Previously, several studies have reported that MHC class II molecules were downregulated in blood monocytes^4,14,18^ in patients with severe COVID-19, however, there was still no consensus on the upregulated genes. We found here that genes related to neutrophil activation, including *S100A8, S100A9* and *S100A12*, were expressed at higher levels in severe COVID-19 patients than those in mild cases. Interestingly, these upregulated and downregulated genes marking monocytic MDSCs whose frequencies are known to increase during various inflammatory conditions^24,25^. Consistent with this immune suppressive status, we also noticed that genes related to cytokine and IFN production were downregulated as well. Thus, contradicted with the inflammatory role by peripheral myeloid cells in severe COVID-19, the loss of mDCs, emergence of MDSC-like monocytes and reduced cytokine production actually suggested a peripheral immune paralysis. Since the MDSC-scores closely associated with lymphopenia and inflammation markers, we speculate a crucial role of MDSCs in dampening immune response and amplifying COVID-19 pathogenesis, which need further functional validation and clinical investigations to evaluate its significance.

The lymphopenia is another prominent feature of immune perturbation of severe COVID-19^2,3,16^. Here, we further revealed that the COVID-19 associated lymphopenia included not only depletion of CD8^+^ T cells, but also significant loss of innate-like T cells, including MAIT, γδ T and NKT cells, similarly reported by another study^13,28^. In contrast, we noticed that the frequencies of CD4^+^ T cells among all CD3^+^ T lymphocytes were actually increased, like Th2, Th17 and Tfh cells. Those CD4^+^ T cell subsets were also more clonally expanded, suggesting their activation status. We suspected that the disturbed T cell compartments may contribute to COVID-19 immunopathogenesis, e.g. the impaired anti-viral responses by innate-like T cells and CD8^+^ T cells, meanwhile amplifying inflammation and inducing aberrant antibody responses by CD4^+^ T cells. Moreover, our transcriptional analysis dispute against the evidence of cytokine production, exhaustion or increased cell death by peripheral T cells in COVID-19 patients noted by several earlier studies^26,29^. Instead, current TCR tracking analysis suggested that the enhanced recruitment of peripheral CD4^+^ and CD8^+^ T cells into lung tissues in COVID-19 patients, where they were induced to made cytokines locally and likely contributed to cytokine storm and peripheral lymphopenia.

In conclusion, we comprehensively delineated the perturbed immune landscapes during SARS-CoV-2 infections from both peripheral blood and infected lungs. These data reveal potential cellular and molecular mechanisms implicated in COVID-19 immunopathogenesis; and identify a peculiar functional dichotomy, with peripheral immune paralysis and broncho-alveolar immune hyperactivation, specifically in severe COVID-19.

## Methods

### Patients

#### Ethics statement

This study was conducted according to the ethical principles of the Declaration of Helsinki. Ethical approval was obtained from the Research Ethics Committee of Shenzhen Third People’s Hospital (2020–207). All participants provided written informed consent for sample collection and subsequent analyses.

Thirteen COVID-19 patients were enrolled in this study at the Shenzhen Third People’s Hospital. Metadata and patients’ samples were collected similarly as previously described^8^: The severity of COVID-19 was categorized to be mild, moderate, severe and critical according to the “Diagnosis and Treatment Protocol of COVID-19 (the 7th Tentative Version)” by National Health Commission of China (http://www.nhc.gov.cn/yzygj/s7653p/202003/46c9294a7dfe4cef80dc7f5912eb1989.shtml). In this study, we grouped patients with mild and moderate COVID-19 as the mild group, and included those with severe and critical diseases as the severe group. Three healthy subjects were enrolled as the control group.

#### qRT-PCR assay for SARS-CoV-2 RNA

In clinical practice, nasal swab, throat swab, sputum, anal swab or BALF could be collected for the SARS-CoV-2 nucleic acid assays. Total nucleic acid was extracted from the samples using the QIAamp RNA Viral kit (Qiagen) and the qRT-PCR was performed using a China Food and Drug Administration-approved commercial kit specific for SARS-CoV-2 detection (GeneoDX). Each qRT– PCR assay provided a threshold cycle (Ct) value. The specimens were considered positive if the Ct value was ≤ 37, and otherwise it was negative. Specimens with a Ct value > 37 were repeated. The specimen was considered positive if the repeat results were the same as the initial result or between 37 and 40. If the repeat Ct was undetectable, the specimen was considered to be negative.

#### Immune cell isolation

For harvesting BALF cells, freshly obtained BALF was placed on ice and processed within 2 hours in BSL-3 laboratory. By passing BALF through a 100 µm nylon cell strainer to filter out cell aggregates and debris, the remaining fluid was centrifuged and the cell pellets were re-suspended in the cooled RPMI 1640 complete medium. For PBMC isolation, immune cells from peripheral blood were isolated by ficoll-hypaque density gradient centrifugation protocol. For subsequent study, the isolated cells were counted in 0.4% trypan blued, centrifuged and re-suspended at the concentration of 2 × 10^6^ /ml.

#### Cytokines measurement by cytometric bead array

Twelve cytokines including IL-1β, IL-6 and IL-8 etc. were detected according to the instruction (Uni-medica, Shenzhen, China, Cat. No. 503022). In brief, the supernatant was taken from BALF after 10 min centrifugation at 1, 000g. Afterwards, 25 µl Sonicate Beads, 25 µl BALF supernatant or plasma, and 25µl of Detection Antibodies were mixed and placed on a shaker at 500 rpm for 2 hours at room temperature. Then 25 µl of SA-PE was added to each tube directly. The tubes were then placed on a shaker at 500 rpm for 30 minutes. The data were obtained by flow cytometry (Canto II, BD) and were analyzed use LEGENDplex v8.0 (VigeneTech Inc.).

#### Flow cytometry

For cell-surface labeling, 1×10^6^ cells were blocked with Fc-block reagent (BD Biosciences). Then, the following antibodies were added and incubated for 30 min, including anti-CD3 (BioLegend, HIT3a), anti-CD14 (BioLegend, 63D3), anti-HLA-DR (BioLegend, L243), and anti-CD45 (BioLegend, 2D1). After incubation, the samples were washed and reconstituted in PBS for flow cytometric analysis on a FACSCanto II flow cytometer.

#### ScRNA-seq library construction

The scRNA-seq libraries were prepared with Chromium Single Cell VDJ Reagent Kits v3 (10x Genomics; PN-1000006, PN-1000014, PN-1000020, PN-1000005) following the manufacturer’s instruction. Briefly, Gel bead in Emulsion (GEM) are generated by combining barcoded Gel Beads, a Master Mix containing 20,000 cells, and Partitioning Oil onto Chromium Chip B. Reverse transcription takes place inside each GEM, after which cDNAs are pooled together for amplification and library construction. The resulting library products consist of Illumina adapters and sample indices, allowing pooling and sequencing of multiple libraries on the next-generation short read sequencer.

#### ScRNA-seq data processing, cell clustering and dimension reduction

We aligned the sequenced reads against GRCh38 human reference genome by Cell Ranger (version 3.1.0, 10x genomics). To remove potential ambient RNAs, we used the remove-background function in CellBender^30^, which removes ambient RNA contamination and random barcode swapping from the raw UMI-based scRNA-seq data. Quality of cells were further assessed by the following criteria: 1) The number of sequenced genes is 200 to 6,000; 2) The total number of UMI per cells is greater than 1,000; 3) The percentage of mitochondrial RNA is less than 15% per cell.

Data integration, cell clustering and dimension reduction were performed by Seurat (version 3)^31^. First, we identified 2,000 highly variable genes (HVGs) which were used for the following analysis using FindVariableFeatures function. Next, we integrated different samples by IntegrateData function, which eliminates technical or batch effect by canonical correlation analysis (CCA). Using those HVGs, we calculate a PCA matrix with the top 50 components by RunPCA function. The cells were then clustered by FindClusters function after building nearest neighbor graph using FindNeighbors function. The cluster-specific marker genes were identified by FindMarkers function using MAST algorithm. The clustered cells were then projected into a two-dimension space for visualization by a non-linear dimensional reduction method RunUMAP in Seurat package.

#### Integrated analysis of peripheral myeloid, NK and T cells

For cells in PBMCs, we integrated the myeloid compartment including mDC and monocytes, or NK and T cells using the similar aforementioned procedure. We re-clustered the myeloid or NK and T cells using the top 20 dimensions of PCA with default parameters. To obtain high resolution cell clusters for each subset, we set the parameter resolution to 1.2. The cell clusters were annotated by canonical markers.

#### Integrated analysis of myeloid, NK and T cells from PBMC and BALF

Myeloid or NK and T cells from PBMC and BALF were integrated separately. For myeloid cells, we extracted macrophage and mDC cells in BALF, and monocyte and mDC in PBMC from the corresponding raw count matrix. The extracted cells were integrated using CCA in Seurat (version 3) as mentioned above. For clustering, the resolution parameter was set to 0.6. Similarly, we extracted NK and T cells in BALF and in PBMC from the corresponding raw count matrix. The extracted cells were integrated using CCA in Seurat (version 3) as mentioned above. For clustering, the resolution parameter was set to 1.5.

#### Analysis of differentially expressed genes

FindMarkers function in Seurat (version 3) with MAST algorithm was used to analyze differentially expressed genes (DEGs). For each pairwise comparison, we run FindMarkers function with parameters of test.use = ‘MAST’. The overlap of differentiated expressed genes among different comparisons were shown by venn diagram. Genes were defined as significant up regulated if average natural logarithm fold change > 0.25 and adjusted P < 0.01. The genes with natural logarithm fold change < –0.25 and adjusted P < 0.01 were considered significantly down regulated. The genes shown on the heatmap having natural logarithm fold change < –0.41 or > 0.41 and adjusted P < 0.01.

#### Gene ontology annotation

ClusterProfiler^32^ in R was used to perform GO term enrichment analysis for the significantly up-regulated and down-regulated genes. Only GO term of Biological Process (BP) was displayed.

#### Calculation of composite score of MHC class II molecules and calprotectin proteins

Composite signature scores of MHC class II molecules and calprotectin proteins of each peripheral CD14^+^ monocytes were calculated using “AddModuleScore” function implemented in the Seurat package. MHC class II score was calculated using the following genes, i.e., *HLA-DMA, HLA-DMB, HLA-DPA1, HLA-DPB1, HLA-DQA1, HLA-DQB1, HLA-DRA, HLA-DRB1*, and *HLA-DRB5*.

Calprotectin protein score was calculated using genes including *S100A1, S100A2, S100A3, S100A4, S100A5, S100A6, S100A7, S100A7A, S100A7L2, S100A7P1, S100A7P2, S100A8, S100A9, S100A10, S100A11, S100A12, S100A13, S100A14, S100A15A, S100A16, S100B, S100G, S100P, and S100Z*. The correlations between MDSC-like scores (“Calprotectin protein score” minus “MHC class II score”) and plasma IL6, CRP levels, and the blood neutrophil / CD3 / CD4 / CD8 cell counts, were calculated using GraphPad Prism 8.4.2. Lines were fitted using the simple linear regression method.

#### Pseudotime trajectory analysis

Trajectories analysis was performed using slingshot^33^ for monocyte-macrophages and T cells separately. For T cells in PBMC, naive CD4 and CD8 T cells were set as the start point for CD4^+^ T cells and CD8^+^ T differentiation trajectory, respectively. For integrated analysis of monocyte-macrophages in PBMC and BALF, we deduced the cell trajectory for each individual using the peripheral monocytes as the start point.

#### Single-cell TCR analysis

The amino acid and nucleotide sequence of TCR chains were assembled and annotated by cellranger vdj function in CellRanger (version 3.1.0). Only cells with paired TCRα and TCRβ chains were included in clonotype analysis. Cells sharing the same TCRα- and TCRβ-CDR3 amino acid sequences were assigned to the same TCR clonotype. We assessed the TCR expansion, TCR transition among cell types and TCR migration between PBMC and BALF using R package STARTRAC (version 0.1.0)

^34^. TCR migration between PBMC and BALF were shown using circos software^35^.

#### Statistics

The Student’s t-test (t.test in R, two-sided, unadjusted for multiple comparisons) was used for pairwise comparisons of the cell proportions between different groups. Statistical difference of TCR expansion index and migration index, between mild and severe disease group, were calculated using t.test in R. Statistical difference of cytokines level between BALF and Blood in Figure 3G were calculated using two-sided wilcox.test in R.

## Data Availability

All data used in this study, including scRNA-seq raw data, expression matrix and scTCR-seq contig annotation that support the findings of this study will be released via a material transfer agreement upon reasonable request.

